# Convalescent plasma therapy for the treatment of patients with COVID-19: Assessment of methods available for antibody detection and their correlation with neutralising antibody levels

**DOI:** 10.1101/2020.05.20.20091694

**Authors:** Heli Harvala, Matthew L Robb, Nick Watkins, Samreen Ijaz, Steven Dicks, Monika Patel, Piyada Supasa, Wanwisa Dejnirattisai, Chang Liu, Juthathip Mongkolsapaya, Abbie Brown, Daniel Bailey, Richard Vipond, Nicholas Grayson, Nigel Temperton, Sunetra Gupta, Rutger J Ploeg, Jai Bolton, Alex Fyfe, Robin Gopal, Peter Simmonds, Gavin Screaton, Craig Thompson, Tim Brooks, Maria Zambon, Gail Miflin, David J Roberts

**Affiliations:** National Microbiology Services, NHS Blood and Transplant, London UK; Statistics and Clinical Studies, NHS Blood and Transplant, Bristol, UK; NHS Blood and Transplant Cambridge, Long Road, Cambridge, CB2 OPT, UK; Virology Reference Department, National Infection Service, Public Health England, Colindale Avenue, London, UK; High Containment Microbiology, National Infection Service, Public Health England, Colindale Avenue, London, UK; Wellcome Centre for Human Genetics, Nuffield Department of Medicine, University of Oxford, Oxford, OX3 7BN, UK; Dengue Hemorrhagic Fever Research Unit, Office for Research and Development, Faculty of Medicine, Siriraj Hospital, Mahidol University, Bangkok, Thailand; Rare & Imported Pathogens Laboratory, Public Health England, Porton Down, UK; Department of Paediatric Medicine, University of Oxford, University of Oxford, Oxford, UK; Medway School of Pharmacy, University of Kent, Chatham, ME4 4BF, UK; Department of Zoology, University of Oxford, Oxford, UK; Nuffield Department of Surgical Sciences, University of Oxford, OX3 9DU, UK; Oxford University Hospitals NHS Foundation Trust, Oxford, OX3 9DU, UK; Nuffield Department of Medicine, University of Oxford, Oxford, UK; NHS Blood and Transplant, North Bristol Park, Filton, Bristol, UK; NHS Blood and Transplant, Oxford, John Radcliffe Hospital, Oxford, OX3 9BQ UK; Radcliffe Department of Medicine and BRC Haematology Theme, University of Oxford, John Radcliffe Hospital, Oxford, OX3 9DU, UK.

**Author notes:** Corresponding author: Dr Heli Harvala, Consultant Medical Virologist, Microbiology Services, NHS Blood and Transplant, Colindale, London, UK.

## Abstract

**Introduction:** The lack of approved specific therapeutic agents to treat COVID-19 associated with SARS coronavirus 2 (SARS-CoV-2) infection has led to the rapid implementation and/or randomised controlled trials of convalescent plasma therapy (CPT) in many countries including the UK. Effective CPT is likely to require high titres of neutralising antibody levels in convalescent donations. Understanding the relationship between functional neutralising antibodies and antibody levels to specific SARS-CoV-2 proteins in scalable assays will be crucial for the success of large-scale collection and use of convalescent plasma. We assessed whether neutralising antibody titres correlated with reactivity in a range of ELISA assays targeting the spike (S) protein, the main target for human immune response.

**Methods:** Blood samples were collected from 52 individuals with a previous laboratory confirmed SARS-CoV-2 infection at least 28 days after symptom resolution. These were assayed for SARS-CoV-2 neutralising antibodies by native virus and lentiviral pseudotype assays, and for antibodies by four different ELISAs measuring antibody binding to different format of viral S proteins. ROC analysis was used to further identify sensitivity and specificity of selected assays to identify samples containing high neutralising antibody levels suitable for clinical use of convalescent plasma.

**Results:** All samples contained SARS-CoV-2 antibodies, whereas neutralising antibody titres of greater than 1:20 were detected in 43 samples (83% of those tested) and >1:100 in 22 samples (42%). The best correlations were observed with EUROimmun IgG ELISA S/CO reactivity (Spearman Rho correlation co-efficient 0.88; p<0.001). Based on ROC analysis, EUROimmun would detect 60% of samples with titres of >1:100 with 100% specificity using a reactivity index of 9.1 (13/22).

**Discussion:** Robust associations between virus neutralising antibody titres and reactivity in several ELISA-based antibody tests demonstrate their possible utility for scaled-up production of convalescent plasma containing potentially therapeutic levels of anti-SARS-CoV-2 neutralising antibodies.

## INTRODUCTION

The emergence of a novel coronavirus as a cause of respiratory disease occasionally leading to severe acute respiratory syndrome (SARS), was first noted in the Hubei province, China in December 2019. From there it rapidly spread to a number of countries including Italy, Iran, Spain and France^1^. Subsequently this virus was classified as SARS coronavirus 2 (SARS-CoV-2) within the genus *Betacoronavirus*^2^ and its associated disease termed COVID-19. Mortality due to COVID-19 is as high as 50% for patients admitted to intensive care units^3^.

The first imported cases of SARS-CoV-2 were identified in the UK at the end of January 2020, and local transmission within UK became evident one month later. As of 1^st^ May 2020, a total of 182,260 cases and 28,131 deaths have been reported and the numbers are predicted to continue to rise in this first pandemic wave. Currently, there are no approved specific antivirals targeting the novel virus and convalescent plasma therapy has been suggested as an immediately available therapy. A systematic review and retrospective meta-analysis including 699 treated patients with SARS-CoV-1 infection or severe influenza, and 568 untreated controls, demonstrated a statistically significant reduction in mortality following treatment in the pooled odds of mortality following treatment, compared with placebo or no therapy (odds ratio 0.25; 95% CI:0.14–0.45)^4^.

Convalescent plasma may be an effective treatment for COVID-19, with success linked to levels of neutralising antibody present in plasma which reduce viral replication and increase viral clearance^5,6^. Virus-specific neutralising antibodies play a key role in viral clearance. The spike (S) protein is responsible for the SARS-CoV-2 attachment and entry to the target cells via the ACE-2 receptor, and neutralising antibodies recognizing the receptor-binding domain (RBD) on the S protein have been shown to block viral entry^7^. Antibodies against other domains of spike protein or possibly even against other proteins may contribute to functional neutralisation of the virus. Neutralising antibodies are known to be detectable in patients approximately 10 to 15 days after the onset of SARS-CoV-2 infection^8^ but this antibody response continues to mature at least for 3 weeks^9^ and potentially longer.

The issue of potential toxicity of convalescent plasma via antibody-dependent enhancement (ADE) also needs to be addressed carefully. It has been shown to occur when non-neutralising or heterotypic antibodies facilitate viral entry into host cells and enhance viral infectivity^10^. It is likely to occur when antibody levels or specificities do not permit neutralisation^11^. For these reasons, it is important to determine neutralising antibody titres in donated plasma as well as a practical cut-off titre level to evaluate not only its safety but also its effectiveness for convalescent plasma transfusion.

Neutralising antibody levels can be either determined directly using native or pseudotype virus in cellular bioassays or be estimated by ELISA if there is an adequate correlation between neutralising antibody titre and ELISA binding reactivity. Neutralising antibody titre can be detected and quantified in a microneutralisation assay format in which samples are assayed for their ability to block infection of cells by SARS-CoV-2. Similarly, a pseudotype assay can be used to measure neutralising antibody levels using a virus construct containing SARS-CoV-2 S protein in the surface of a luciferase tagged vesicular stomatitis virus or lentivirus viral vector^12,13^. Both types of assays use suitably characterised target cells. Whereas a limitation of neutralisation assays using live virus is the necessity to undertake work at BSL-3 laboratory, pseudotype assay is more suitable for high-throughput screening of convalescent plasma donors as it can be done at BSL-2 facility.

In the current study, we have first determined the neutralising antibody levels in our convalescent plasma donors and estimated a cut-off to be used in clinical trials. Secondly, we have also assessed whether there is a correlation between neutralisation antibody titres (measured either using native SARS-CoV-2 or pseudotype assay) and ELISA reactivity using a variety of assays formats including (1) SARS-CoV-2 infected cell lysate, (2) recombinant S protein assay and two commercial ELISAs. Identification of a suitable high throughput assay is required urgently to support scaling up convalescent plasma production and to support the comparison of data between countries.

## MATERIALS AND METHODS

### Convalescent plasma donors

We initiated the collection of convalescent plasma using the established infrastructure and standard UK donor selection guidelines during March 2020; serum and EDTA blood samples were collected from individuals with a previous laboratory confirmed SARS-CoV-2 infection at least 28 days after the resolution of their symptoms. These donors samples were submitted to Public Health England and tested initially for SARS-CoV-2 RNA by RT-PCR assay^14^ as well as SARS-CoV-2 antibodies using a native virus antigen ELISA and microneutralisation assays both based on the UK prototype strain (GISAID accession number EPI/ISL/407073), and then subsequently subjected to testing by pseudotype neutralisation assay and trimeric spike ELISA. Basic donor information including age, gender and virology testing data were collected.

### Ethical statement

Signed donor consent is collected from each donor at the time of donation, including convalescent plasma donors and covers all the testing including anti-SARS-CoV-2 antibody testing. It also covers holding information about them including their health, attendances and donations and using their information for the purposes explained in the donor welcome booklet and data protection leaflet which donors are asked to read at the time of donation. This includes using data for the purposes of clinical audit to assess and improve the service and for research, specifically to improve our knowledge of the donor population. Consent is collected using NHS Blood and Transplant approved consent forms and these follow Blood Safety and Quality Regulations enforced by the Medicines & Healthcare products Regulatory Agency. The study was performed under NHS Health Research Authority guidance, and approved by the NHSBT Blood Supply Clinical Governance Committee.

### Infected virus lysate assay

Native virus antigen ELISA was modified from a previously described MERS-CoV assay^15^. Microplate bound detergent extracted lysates of SARS-CoV-2 (isolate England/02/2020) infected Vero E6 cells and uninfected cells were reacted with a serial dilution of convalescent plasma obtained from recovered patients with a previous laboratory confirmed SARS-CoV-2 infection in an indirect ELISA format. Virus lysates contain a mixture of viral proteins expressed in Vero E6 cells, including viral nucleocapsid and S proteins. ELISA index value was defined as the difference between infected and uninfected cell reactivity expressed relative to control calibrator serum.

### Microneutralisation assay and neutralising antibody titre

SARS-CoV-2 (isolate England/02/2020) specific neutralising antibody levels were measured using a modification of the WHO influenza microneutralisation methodology^16^. Briefly, virus was incubated with a serial dilution of convalescent plasma obtained from recovered patients, after which a suspension of Vero E6 cells were added. After 22 hours cells were fixed and in-cell SARS-CoV-2 nucleoprotein (NP) expression determined by ELISA. The virus neutralising antibody titre was determined as the serum concentration that that inhibited 50% SARS-CoV-2 NP expression. All work was undertaken in a BSL-3 laboratory.

### Enzyme-linked trimeric spike immunosorbent assay (ELISA - Oxford)

Antibodies to the trimeric spike (S; based on YP009724390.1) protein were detected by ELISA as previously described, using 2% skimmed milk in PBS as a blocking agent and ALP-conjugated anti-human IgG (A95455; Sigma) at 1:10,000 dilution^12^. Optical densities (ODs) were measured at 405nm.

### Pseudoparticle neutralisation test

A lentivirus-based SARS-CoV-2 pseudoparticle assay was performed as previously described (accession number: YP009724390.1)^12^. Neutralisation was measured by the reduction in luciferase gene expression. The 50% inhibitory dilution (EC_50_) was defined as the plasma dilution at which the relative light units (RLUs) were reduced by 50% compared with the virus control wells after subtraction of the background RLUs in the groups with cells only.

### Commercial assays, EUROimmun (IgG) and Fortress (total antibodies)

EUROimmun assay is based on S1 protein, and Fortress assay on RBD of S protein. These assays were performed according to the manufacturer’s recommendation (EUROimmun, PerkinElmer, London, UK and Fortress Diagnostics, Belfast, Northern Ireland).

### Statistics

Associations between test assays were compared using Pearson correlation coefficients and the non-parametric Spearman’s rank correlation. P-values were derived using Student’s t test for correlations and Pearson correlation coefficient, under the null hypothesis that the correlation was zero. The sensitivity and specificity were calculated to assess the performance of the different assays in classifying the level of neutralising antibody titres obtained by microneutralisation assay using live SARS-CoV-2 virus. Exact binomial confidence intervals were used to derive confidence intervals.

## RESULTS

The initial assessment included samples from 52 recovered patients who would qualify as donors of convalescent plasma for clinical trials. They were all males and at least 28 days from the recovery after laboratory confirmed SARS-CoV-2 infection. They were sampled during the first two weeks of April, implying that their illness was at the beginning of March or earlier. Therefore, they would all have been hospitalised as a part of containment strategy. However, no data on severity of their infection is currently available. EDTA and serum samples were obtained from each and a whole blood donation was collected from 10. All samples were submitted to Public Health England Colindale and distributed from there to the University of Oxford and Public Health England Porton Down for further testing. All samples tested negative for SARS-CoV-2 RNA. Assay specificity (particularly the rate of false reactives) has not been included in this analysis.

SARS-CoV-2 neutralising antibodies were detected by 43 out of 52 tested samples using a cut-off titre 1:20; the highest detectable titre was 1:4096 (Figure 1). In other assays, SARS-CoV-2 antibodies were detected in most samples tested by pseudotype assay (47/51), lysate ELISA (47/50), EUROimmune (47/50) and in all samples by trimeric spike ELISA (51/51) and Fortress total antibody ELISA (50/50). Based on these initial observations, all assays demonstrated good sensitivity for detecting antibodies in the study subjects from 28 days after their recovery. For most assays, quantitative measures of serological reactivity (IC_50_ in the pseudotype assay, optical densities (ODs) or signal to cut-off ratios (S/CO)) showed associations with neutralising antibody titres based on the live virus microneutralisation assay (Fig. 1).

**Figure 1.**
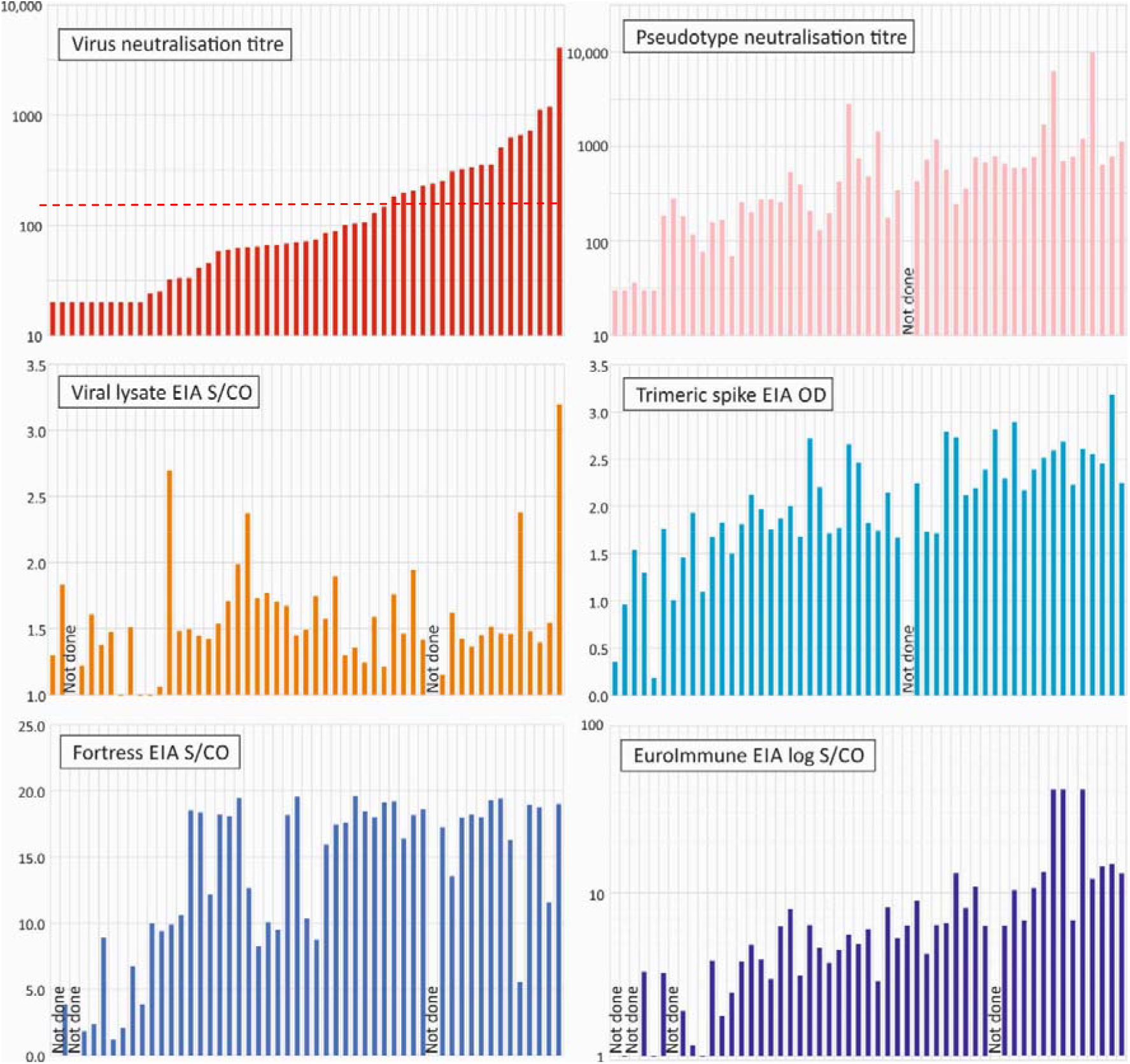
Comparison of neutralising antibody titres with reactivity in other assays. Comparison of neutralising antibody titres of the 52 test samples in the virus neutralisation assay with those of the pseudotype assay and reactivities in EIAs. In all graphs, samples were ordered by virus neutralising antibody titres. The following assay cut-off values were used: 0.049 for trimeric spike EIA, 1.0 for Fortress EIA, and 1.1 for EUROimmun.

We have further assessed the correlation between neutralising antibody titre and serological reactivities in different ELISA platforms (Fig. 2) where Pearson correlation coefficients and the non-parametric Spearman’s Rank correlation tests were performed. The Pearson correlation tests for a linear association between variables (using log transformed values for the neutralisation, pseudotype and Euroimmun assays; R^2^ values) whereas the Spearman’s coefficient determined correlations in ranking irrespective of magnitude. A further comprehensive of pairwise comparison between all assays is provided in Supplementary Data (Fig. S1).

**Figure 2.**
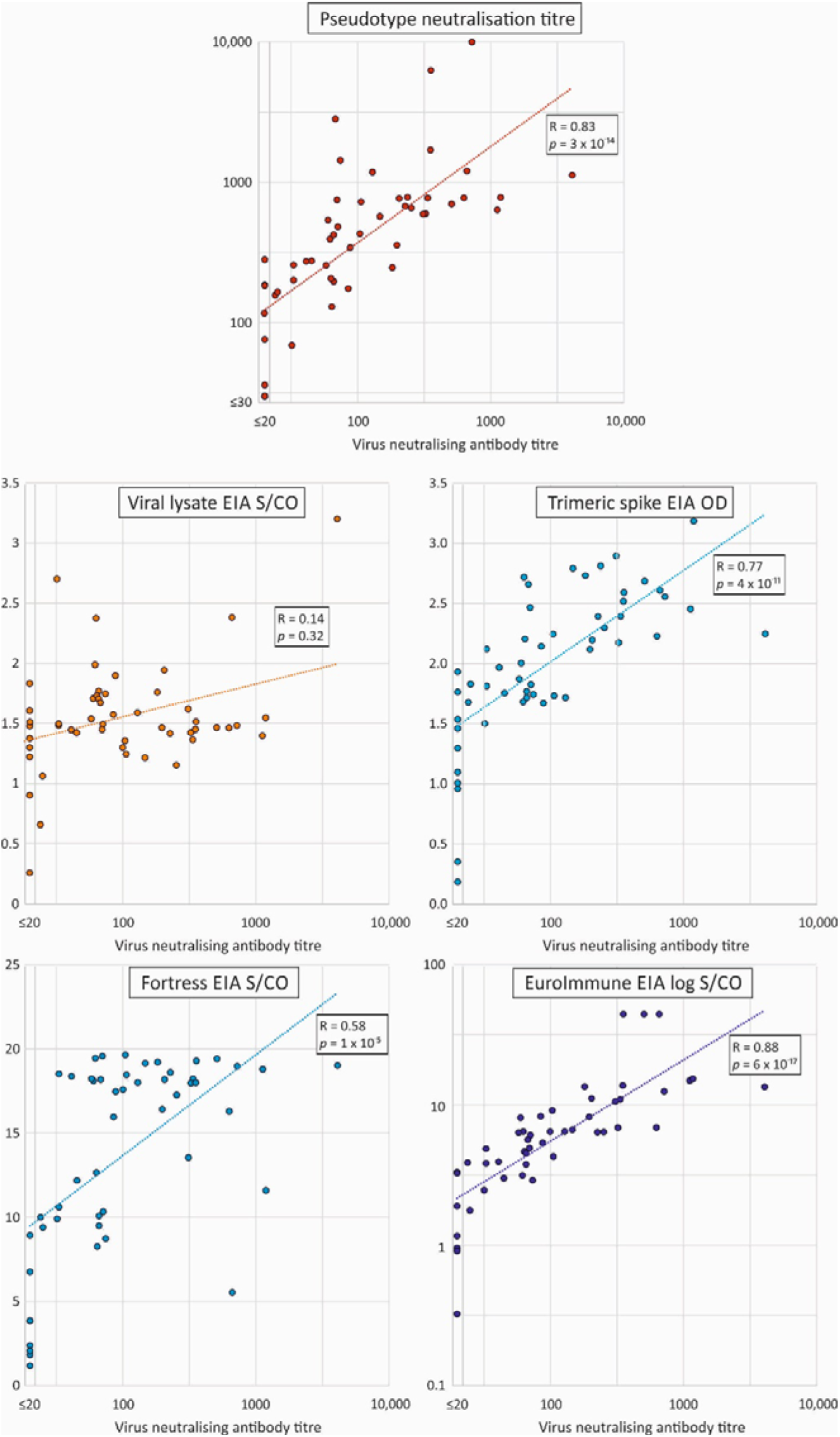
Correlations between neutralising and pseudotype antibody titres and reactivities in EIAs. **S**catter plots of neutralising antibody titres of test samples in the virus neutralisation assay with those of the pseudotype assay and reactivities in EIAs. A line of best fit was estimated by linear regression usinglog transformed values for the virus and pseudotype neutralising antibody assays and the Euroimmun EIA. Correlation coefficients and (two-tailed) *p* values were calculated by Spearman non-parametric test.

The strongest correlation was observed between neutralising antibody titres and reactivity in the EUROimmun IgG ELISA (Spearman’s rank correlation: 0.88; p<0.0001, n=48). Correlations were also observed between neutralising antibody titres with IC50 values in the pseudotype assay (Spearman’s rank correlation: 0.82; p<0.0001, n=51) and trimeric spike ELISA (Spearman’s rank correlation: 0.76; p<0.0001, n=51).

The predictive value of pseudotype IC50 values and reactivity in the EUROimmun assay for neutralising antibody titres was determined by ROC analysis (Fig. S2; Table S1; Suppl. Data). In the analysis, a neutralising antibody titre of 1: 100 was selected as this represents a likely therapeutic threshold for plasma donation selection (see discussion). Five potential cut-off values in the EUROimmun ELISA (S/CO values between 6.37 – 10) were investigated for practicality and sensitivity; a value of 9.1 identified 65% of donations above the 1:100 neutralising antibody threshold with no false identification of donations below this neutralising antibody threshold. In contrast, the pseudotype assay was unable to identify 50% or more donations >1:100 without false identification.

## DISCUSSION

Here, we have described the first evaluation of the relationship between neutralising antibody titres and measures of antibodies to SARS-CoV-2 proteins in a variety of assays. These data can guide selection of units of convalescent plasma for clinical use and for randomised clinical trials.

Our initial observation of convalescent plasma donors sampled at least 28 days after the recovery from a laboratory confirmed SARS-CoV-2 infection, showed all of them demonstrated serological evidence of past SARS-CoV-2 infection in one or more assays whereas the SARS-CoV-2 neutralising antibody levels detected varied from low (1:20) to high (1:4096; Figure 1). Furthermore, approximately 43% of donor samples showed neutralising antibody titres of greater than 1:100. These neutralising antibody titres correlated with values obtained by pseudovirus assay; a titre 1:100 corresponded to 1:300 calculated based on luminescence reading. The pseudotype assay can be automated and does not require working with live virus in a biosafety level 3 laboratory and therefore provides a potential approach to directly measuring neutralising antibody levels at scale.

In a previous study, most convalescent plasma donors with previous COVID-19 showed high neutralising antibody titres of at least 1:160 determined by plaque reduction neutralisation test (PRNT; 39/40). For convalescent plasma therapy, only donations with antibody titres above 1:640 were used^5^. In a separate study, donations with a neutralising antibody titre equal or higher than 1:80 based on microneutralisation test were used successfully^6^. It is important to note that antibody titres obtained by different assays may not be comparable; based on previous data on SARS-CoV-2 neutralising antibody titres obtained by PRNT were approximately 4-fold higher than those obtained by a CPE based microneutralisation assay^17^. Further comparative work is required to determine how the neutralising antibody level compares with the PRNT titres and also with assays performed outside the UK. The future availability of international standards will facilitate such comparisons.

A minimum neutralising antibody titre in convalescent plasma needs to be determined before plasma is supplied for clinical trials. This needs to be balanced with the difficulty of collecting a required number of such components while providing a sufficient dose of antibodies to potentially be effective. For the planned trial, the use of plasma with a too low cut-off may prevent or prolong a clear demonstration of efficacy; conversely a too high cutoff may prevent a sufficient supply of plasma to fulfil trial needs. The chosen neutralising antibody level 1:100 was selected as a pragmatic cut-off that enables an estimated 40 % of collected plasma to be used. The actual dose of neutralising antibody given to patients also depends on the number of units given and giving two units from different donors may substantially increase the mean dose to more than 1:300. Although considered potentially effective, how this level compares with that used in previous studies requires further work. This cut-off will be reviewed after a larger number of samples have been analysed to see if supply is meeting demand.

In order to support the scaling up the convalescent plasma production, it is important to identify a suitable high throughput ELISA assay which can be used to estimate the neutralising antibody levels in convalescent plasma samples and hence could determine which donations are offered for a clinical use. Serological reactivity in both the EUROimmun SARS-CoV-2 IgG ELISA and the trimeric spike SARS-CoV-2 ELISA showed a strong correlation with neutralising antibodies obtained either by live virus or by pseudotype assay. Although the EUROimmun assay has been shown to lack sensitivity for samples collected from patients with recent infection^18^, we have shown that it could be used to identify donations containing high levels of neutralising antibodies with a good level of specificity. By selecting a S/CO cut-off value of 9.1, the assay would only identify units were neutralising antibody titre was 1:100 or higher. This is consistent with a previous finding where plasma with high titers of neutralising SARS-CoV-2 antibodies showed also higher titres of RBD, spike domain 1 or 2, specific binding antibodies^8^. Trimeric spike ELISA contains RBD domain, whereas EUROimmun is based on spike domain 1. However, it is important to note that this is based on testing a pre-selected cohort of individuals at least 28 days after the recovery from a previous laboratory confirmed SARS-CoV-2 infection. The evaluation should be repeated if these criteria are changed.

As only a relatively small number of samples from convalescent plasma donors have been tested so far, we propose that several assay formats should be employed in a larger group of donors to validate these findings before the scaling up can be finalised. Nevertheless, the results provide guidance for the many convalescent plasma programmes in progress around the world.

Neutralising antibody levels are dependent in part on the timing of collection relative to the recovery from infection. Seroconversion following SARS-CoV-2 infection has been observed between 8 and 21 days after the onset of symptoms ^9,19-21^, and higher levels of antibodies have been determined in plasma collected at least 14 days after the symptom resolution^5^. It is likely that the antibody maturation continues for longer as demonstrated for other viruses, and hence the collection point 28 days after recovery has been chosen here. This maximises the chances of collecting the most clinically effective donations. However, it is still unclear how long neutralising antibody levels are maintained and hence repeat testing will be performed at every donation.

Higher neutralising SARS-CoV-2 antibody levels have been associated with the older age and a worse clinical outcome^8,21^ although good neutralising antibody levels have also been measured from individual patients with milder infections^22,23^. The monitoring of neutralising antibody levels in different patient groups (including females not included in this study) and over time is required and will inform future screening strategies.

In conclusion, we have demonstrated here a correlation between the neutralising antibody level and antibody reactivity measured by ELISA which will allow scaling-up the convalescent plasma production. However, the continuous monitoring of assay performance, antibody decay and adaptation of selection strategies will be required in order to deliver the best clinical outcomes for patients receiving neutralising SARS-CoV-2 antibodies containing convalescent plasma therapy.

## Data Availability

All data used in the study is available from the corresponding author on request

## ACKNOWLEGDEMENTS

We would like to thank all to donors who have kindly donated convalescent plasma. We are grateful for everybody within NHS Blood and Transplant who have participated to the Convalescent Plasma programme, and at the operations level to the donor outreach, collection, transporting, processing and storing convalescent plasma. We would also like to thank the staff members of the Virus Reference Division at Public Health England Colindale.

## AUTHORS’ CONTRIBUTIONS

Heli Harvala, Nick Watkins, Samreen Ijaz, Steven Dicks, Monika Patel, Piyada Supasa, Wanwisa Dejnirattisai, Chang Liu, Juthathip Mongkolsapaya, Abbie Brown, Daniel Bailey, Richard Vipond, Nicholas Grayson, Nigel Temperton, Craig Thompson and Robin Gopal performed the sample acquisition, laboratory testing and reporting of the pseudotype and ELISA testing. Matthew L Robb designed the data collection pathways and statistical analysis of the laboratory findings. Heli Harvala, Nick Watkins, Sunetra Gupta, Rutger J Ploeg, Jai Bolton, Alex Fyfe, Peter Simmonds, Gavin Screaton, Tim Brooks, Maria Zambon, Gail Miflin and David J Roberts conceived and designed the specifics of the study, the data interpretation and drafting of the manuscript. All co-authors contributed to the editing and final drafting of the manuscript and figures.

## CONFLICT OF INTEREST STATEMENTS

The authors declare no conflict of interest

## ROLE OF FUNDING SOURCE

The funders played no role in the design, execution or reporting of the study.

## ETHICS COMMITTEE APPROVAL

Signed donor consent is collected from each donor at the time of donation, including convalescent plasma donors and covers all the testing including anti-SARS-CoV-2 antibody testing. It also covers holding information about them including their health, attendances and donations and using their information for the purposes explained in the donor welcome booklet and data protection leaflet which donors are asked to read at the time of donation. This includes using data for the purposes of clinical audit to assess and improve the service and for research, specifically to improve our knowledge of the donor population. Consent is collected using NHS Blood and Transplant approved consent forms and these follow Blood Safety and Quality Regulations enforced by the Medicines & Healthcare products Regulatory Agency.

## FUNDING

This work was supported by the Medical Research Council [grant number MC_PC_19059]. National Institute for Health Research Biomedical Research Centre Funding Scheme (to G.R.S.). PS and G.R.S. are supported as a Wellcome Trust Senior Investigators (grant 095541/A/11/Z; WT109965/MA). NG was supported via grant to Philip Goulder (WTIA Grant WT104748MA) and a grant to John Frater (Medical Research Council MR/L006588/1). CPT and SG are funded by an ERC research grant ‘UNIFLUVAC’ and two MRC CiC grants (Ref: BR00140).

---

**Table 1.** Threshold values for optimal sensitivity and specificity of Euroimmun and pseudotype neutralisation assays by ROC analysis.

| **Cut-off value** | **Sensitivity (95% CI)** | **Specificity (95% CI)** |
| --- | --- | --- |
| <u>*Euroimmun S/CO*</u> |  |  |
| 6.37 | 0.95 (0.76, 1.00) | 0.89 (0.98, 0.77) |
| 6.64 | 0.76 (0.53, 0.92) | 0.93 (1.00, 0.83) |
| 8.19 | 0.68 (0.48, 0.83) | 0.96 (0.99, 0.85) |
| **9.1<sup>1</sup>** | 0.65 (0.45, 0.81) | 1.00 (1.00, 0.92) |
| 10 | 0.52 (0.30, 0.74) | 1.00 (1.00, 1.00) |
| <u>*Pseudotype neut. titre*</u> |  |  |
| 573 | 0.86 (0.64, 0.97) | 0.90 (0.98, 0.73) |
| 770 | 0.48 (0.26, 0.70) | 0.93 (0.99, 0.78) |

<sup>1</sup>Optimal value selected for donation selection shown in bold.

**Figure 3.**
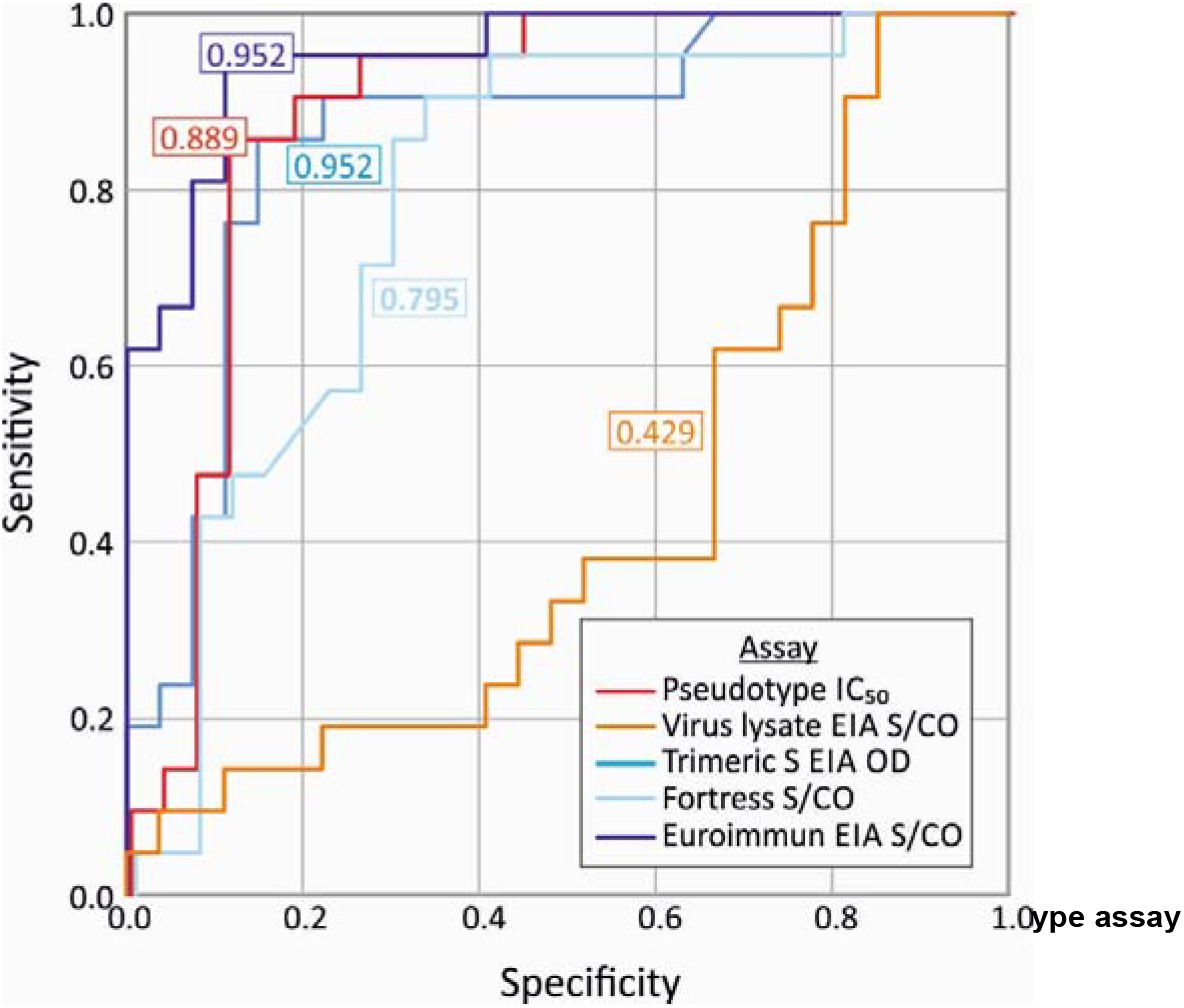
ROC analysis of serology assays predicting virus neutralising antibody titres of ≥1/100. OC curves for the pseudotype, virus lysate and three EIAs to correctly identify samples with neutralising antibody titres of 1:100 and over in the virus neutralisation assay (n=48). Areas under the curve for each assay shown in colour-coded boxes.

## Notes

### Competing Interest Statement

The authors have declared no competing interest.

